# Open Development and Clinical Validation Of Multiple 3D-Printed Sample-Collection Swabs: Rapid Resolution of a Critical COVID-19 Testing Bottleneck

**DOI:** 10.1101/2020.04.14.20065094

**Authors:** Cody J Callahan, Rose Lee, Katelyn E. Zulauf, Lauren Tamburello, Kenneth P. Smith, Joe Previtera, Annie Cheng, Alex Green, Ahmed Abdul Azim, Amanda Yano, Nancy Doraiswami, James E. Kirby, Ramy A. Arnaout

## Abstract

The SARS-CoV-2 pandemic has caused a severe international shortage of the nasopharyngeal swabs that are required for collection of optimal specimens, creating a critical bottleneck in the way of high-sensitivity virological testing for COVID-19. To address this crisis, we designed and executed an innovative, radically cooperative, rapid-response translational-research program that brought together healthcare workers, manufacturers, and scientists to emergently develop and clinically validate new swabs for immediate mass production by 3D printing. We performed a rigorous multi-step preclinical evaluation on 160 swab designs and 48 materials from 24 companies, laboratories, and individuals, and shared results and other feedback via a public data repository (http://github.com/rarnaout/Covidswab/). We validated four prototypes through an institutional review board (IRB)-approved clinical trial that involved 276 outpatient volunteers who presented to our hospital’s drive-through testing center with symptoms suspicious for COVID-19. Each participant was swabbed with a reference swab (the control) and a prototype, and SARS-CoV-2 reverse-transcriptase polymerase chain reaction (RT-PCR) results were compared. All prototypes displayed excellent concordance with the control (κ=0.85-0.89). Cycle-threshold (Ct) values were not significantly different between each prototype and the control, supporting the new swabs’ non-inferiority (Mann-Whitney U [MWU] p>0.05). Study staff preferred one of the prototypes over the others and the control swab overall. The total time elapsed between identification of the problem and validation of the first prototype was 22 days. Contact information for ordering can be found at http://printedswabs.org. Our experience holds lessons for the rapid development, validation, and deployment of new technology for this pandemic and beyond.

## Introduction

Since the emergence of the COVID-19 pandemic, more than 2.5 million cases have been diagnosed worldwide (*1*). These diagnoses were made using material collected from NP swabs, which provide the highest sensitivity for detecting SARS-CoV-2 infection during early infection using commercial RT-PCR-based assays. An NP swab is an FDA Class I exempt medical device roughly 15 cm in length and 2-3mm in diameter designed to collect secretions from the posterior nasopharynx (Figs. 1a, left and 1b, top). The head of the swab is generally coated with short synthetic filaments called flock. The swab is inserted into the nasopharynx, rotated several times to collect material, and then placed into a vial containing a few milliliters of transport media. A breakpoint on the shaft enables detachment and release of the head into the vial, which is then sealed and sent for testing.

**Figure 1:**
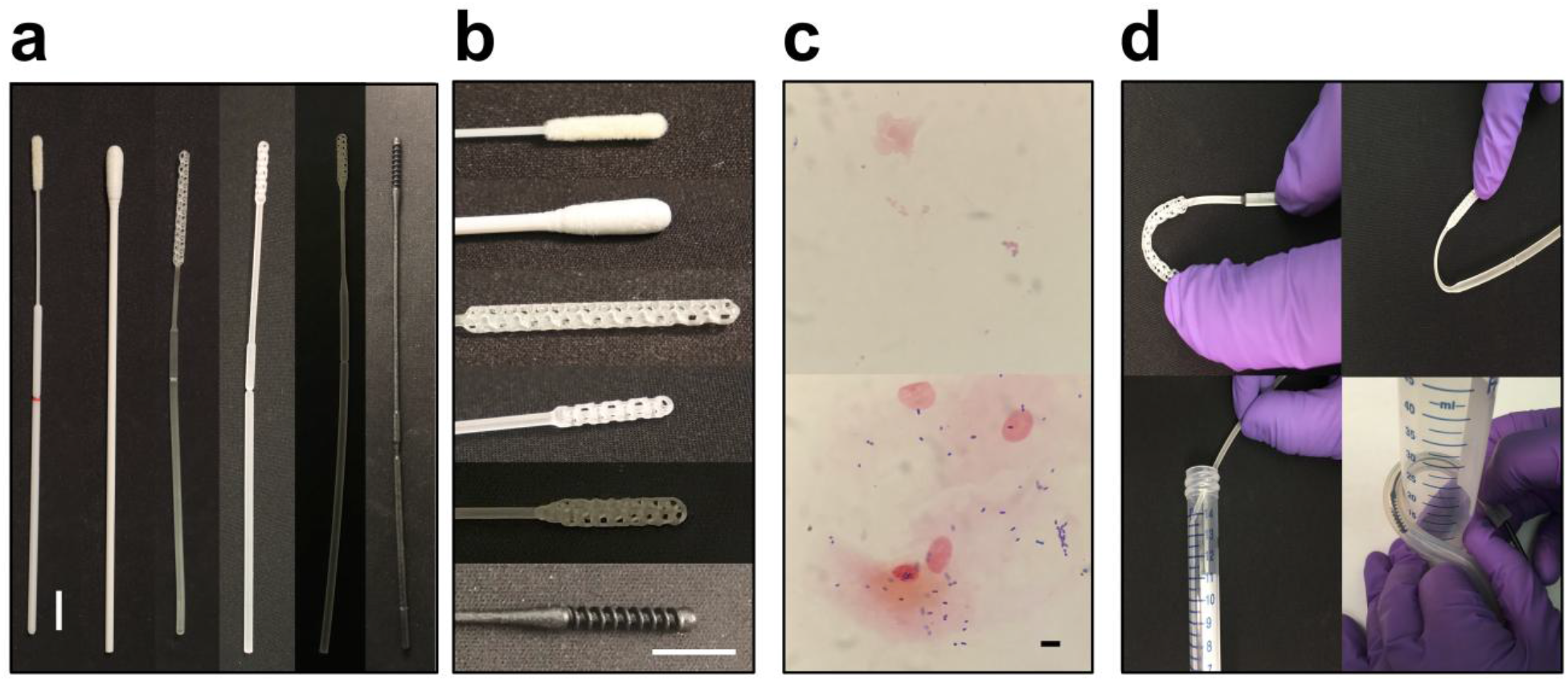
Control and prototype swabs. **(a)** From left to right: the control swab (Copan 501CS01), a repurposed urogenital cleaning swab approved for NP testing through our process, Prototype 1 (Resolution Medical), Prototype 2 (EnvisionTec), Prototype 3 (Origin.io), and Prototype 4 (Hewlett Packard). **(b)** From top to bottom, closeups of the heads of the swabs in (a). Scale bars, 1cm. **(c)** Examples of Gram stain of cheek swab using control (top) and prototype swabs. Scale bar, 10µm. **(d)** Examples of materials testing. Clockwise from top left: head flexibility and robustness to fracture, neck flexibility and robustness to fracture, robustness to repeat insertion into and removal from a tortuous canal (diameter 3cm), and breakpoint evaluation.

The rapid spread of SARS-CoV-2 has resulted in severe shortages of NP swabs, due to both manufacturing stoppages resulting in decreased supply and the spread of the pandemic resulting in unprecedented demand (*2*). To address the swab shortage, hospitals and other testing centers have repurposed other commercially available swabs (e.g. throat, urogenital) to collect nasal epithelial mucosa for testing (Fig. 1a, second from left and 1b, second from top). However, such swabs are suboptimal for swabbing the nasopharynx due to differences in size and flexibility and the possibility they contain PCR-inhibitory materials (*3, 4*). Material from other sites has not yet been shown to be able to substitute for swabbing the nasopharynx.

One solution to the swab crisis is to design and 3D-print swabs. Advantages of 3D printing include simplicity (avoiding the multistep process of applying flock), the widespread availability of 3D printing capacity, and the ability to iterate prototypes rapidly (*5*). To resolve the swab-shortage crisis, we have been coordinating an open collaborative process that has brought together many medical centers, individuals, academic laboratories, and both new and well established manufacturers (*6*). As part of this process, we have been testing and continuously providing feedback on prototype swabs in order to proceed rapidly but safely toward the development of swabs that can be used clinically, at volumes equal to the need. The openness of the process was a conscious decision supported by a substantial body of scientific literature, including the previous experience of the present authors, that demonstrates the advantages of openness over closed or hybrid approaches (*7–9*). At our institution, this process has led to an ongoing clinical trial of several prototype swabs, the first results of which we report here.

## Materials and Methods

### Process

We created a public repository using GitHub, a free website most often used by programmers to co-develop computer code (http://www.github.com/rarnaout/Covidswab)^6^. We provided a clear description of the problem and updated the repository with whatever we learned and encouraged others to do the same. By tapping our personal and professional networks, we nucleated an ad hoc network of manufacturers that included companies, academic groups, and individuals. This grew to include other medical centers interested in helping develop and test new swabs. These other groups were given the ability to add to the repository as desired.

We devised a three-phase process consisting of preclinical evaluation (Phase I), production considerations (Phase II), and field testing (Phase III). We described these processes on the repository for all to see. We took high-resolution photographs of all prototypes and stored Phase I results in a Microsoft Excel (Microsoft Corporation, Redmond, WA, USA) spreadsheet that remains publicly available in the repository. All contributors could see each others’ designs and our feedback and iterated accordingly.

We made our personal contact information freely available to facilitate communication and speed the delivery of prototypes. We involved representatives of our institution’s nursing, legal, intellectual property, leadership, purchasing, human resources, communications, and contracting teams and the institutional review board early and often in order to facilitate open development, reassign idled staff to our process, and minimize lead times during the rapidly changing situation.

### Phase I: Preclinical evaluation

#### Design

An infectious disease physician, clinical pathologist (clinical microbiologist), and respiratory therapist tested each prototype swab for design and mechanical properties (Fig. 1c-d). These included size measurements of the head, neck, shaft, and breakpoint (requirement of ~15cm to reach the posterior nasopharynx; head diameter of 1-3.2mm to pass into the mid-inferior portion of the inferior turbinate and be able maneuver appropriately without catching on anatomical variants such as septal spurs or a deviated nasal septum); surface properties such as smoothness (with roughness leading to an unpleasant feel and risk of bleeding); flexibility vs. brittleness of the head, neck, shaft, and breakpoint (to avoid fracture during use); durability (e.g. ability to tolerate 20 rough repeated insertions into a 4-mm-inner-diameter clear plastic tube curved back on itself with a curve radius of ~3 centimeters; ability to bend tip and neck 90 degrees without breaking; ability to restore to initial form following bend of 45 degrees; Fig. 1d); strength (resist breakage under rough but reasonable manipulation); and other factors as applicable (e.g. stickiness, smell).

#### Collection sufficiency

We assessed the ability to collect sufficient material for testing using Gram stain of a swab of the interior cheek smeared onto a standard microscopy slide as a surrogate for NP swabbing and comparison to Gram stain of a swab of the interior cheek using Copan Diagnostics, Inc. (Mantua, Italy) model 501CS01 NP swab as the control (Fig. 1c). Slides were heat fixed and Gram stained according to the BD BBL gram stain test kit protocol (*10*). Slides were examined at 40x magnification for the presence of both epithelial cells and bacteria. Prototypes were passed if they collected a comparable quantity of the material as the control.

#### PCR compatibility

We tested PCR compatibility by incubating the head overnight in 3 mL of modified CDC VTM (Hank’s balanced salt solution containing: 2% heat inactivated FBS, 100μg/mL gentamicin, 0.5μg/mL fungizone, and 10mg/L Phenol red (*11*)) to allow any PCR-inhibitory material to leach into the medium, spiking 1.5mL with 200 copies/mL of control SARSCoV-2 amplicon target (representing 2 times the limit of detection on our system), vortexing, and testing using the Abbott RealTime SARS-CoV-2 Assay on an Abbott m2000 RealTime System platform (*12*), following the same protocol as for clinical testing. PCR-positive prototypes passed.

#### Phase II: Production considerations

We considered stability to autoclaving by repeating Phase I testing on post-autoclaved materials; manufacturers’ short-term strategies for individual packaging; and manufacturers’ stated ability to produce at least 10,000 swabs per day (at the time roughly a week’s worth of swabs for a mid-sized testing center) within a week’s notice. We considered differences in supply chain to minimize the risk of future crises.

### Phase III: Field testing

#### Trial design and oversight

COVIDSwab is an adaptive trial for evaluating the performance of prototypes compared to the control (see above). Participants under clinical suspicion for COVID-19 who were scheduled for standard clinical SARS-CoV-2 RT-PCR testing with a control swab were asked also to be swabbed afterward with a single prototype. Prototypes were collected and tested until at least 10 positive and 10 negative results on control swabs were obtained (*13*). Sample collection was performed by trained nursing or respiratory-therapy staff (“study staff”) overseen by the respiratory therapy department at BIDMC. The Clinical Microbiology laboratories oversaw data collection. This study was reviewed and approved by the institutional review board of Beth Israel Deaconess Medical Center (protocol number 2020P000323).

#### Participants

Participants were individuals clinically suspected of COVID-19 who were brought to the drive-through/walk-up (“drive-through”) COVID-19 testing site at BIDMC. Adults over 18 years of age were given a participant information sheet by study staff and asked whether they would agree to being swabbed with a prototype swab performed by a trained nurse or respiratory therapist in addition to the control swab required for testing. Individuals with known thrombocytopenia of <50,000 platelets/µl were excluded from the study to avoid risk of mild bleeding.

#### Trial procedures

Prototype swabs were individually packaged and autoclaved at BIDMC for sterilization according to manufacturer protocols. Swabbing was performed per standard protocol. Participants were first swabbed with the control swab, then the prototype. Choice of naris for each swab was left to study staff and the participant. Approximately half of all drive-through arrivals participated. Control and prototype swabs were placed in separate vials of VTM and transported to the BIDMC Clinical Microbiology Laboratories where each sample was tested on the Abbott m2000 SARS-CoV-2 RT-PCR platform as per standard clinical protocol.

#### Statistical analyses

RT-PCR results are reported categorically as either positive or negative. We tested categorical concordance using Cohen’s kappa (*14*). For each positive test, the Ct value (the RT-PCR cycle number at which the sample first turns positive) was obtained from the Clinical Microbiology Laboratories. Higher values reflect lower viral load in the sample.

We tested for systematic bias in Ct values by comparing values for controls vs. prototypes using MWU (*15*). This tested the null hypothesis that values for controls and prototypes are drawn from the same underlying distribution; *p*>0.05 was interpreted as no bias. For discordant (positive control/negative prototype or vice versa) samples, the negative was assigned a Ct value of 37, the total number of cycles run. As a second test for bias, we compared (again by MWU) the distribution of differences in Ct values between control and prototype swabs to the distribution of differences between two control swabs taken within 24 hours (quality-control data independent of our study). This tested the null hypothesis that the differences between control and prototype swabs and the differences between two control swabs are drawn from the same underlying distribution; *p*>0.05 was interpreted as no bias.

To quantify relative preferences among the prototypes, we gave study staff members printouts of all six possible pairs of swabs (a “round robin”), in randomized order, and for each pair asked them to circle their preference (A-B testing). We collated the results and assessed preferences.

## Results

### Open process

In the first days of the development effort GitHub repository (*6*) was established to serve a public resource and knowledge base. We updated the repository continuously with design information and test results. These updates included high-resolution images of prototypes submitted to us for testing (*6*), a public database of results of our Phase I testing, and periodic updates and guidance based on our experiences. Open communication facilitated rapid design iteration by providing anyone interested with a way to quickly understand the required specifications and to learn from each other’s experiences.

### Phase I testing

To date we have evaluated 48 materials and 160 designs submitted to us for testing by 4 individuals, 2 laboratories, and 18 companies, for a total of 24 manufacturers. Seven (4.4%) have passed Phase I testing. Most failures were either for inappropriate materials, including some that were sticky or brittle, or for inappropriate designs, including those with sharp heads. Prototypes from 19 manufacturers went through at least two iterations, with a maximum of 28 prototypes from one manufacturer (Prototype 4 below; Fig. 1). The rate-limiting steps were receipt of new prototypes, with slow mail delivery during the pandemic being a major contributor, and PCR-compatibility testing, as testing patient samples took priority over testing prototypes. Communication with and responsiveness by manufacturers were considered outstanding.

### Phase II and III prototypes

Four prototypes passed Phase II testing, all of which have completed our Phase III clinical trial: these are prototypes from the 3D-printing manufacturers Resolution Medical (with technology from Carbon3D), EnvisionTec, Origin.io, and HP Inc. (Prototypes 1-4, respectively; Fig. 1a). Like control swabs, the prototypes were 15-16cm in length with 1-3cm length radially symmetric heads 2-3mm in diameter, a thin neck 4-7cm long and 1-2mm in diameter, and a thicker shaft 2-4mm in diameter, with a breakpoint most often 7-8cm from the tip of the head. The materials were plastics and resins such as Keysplint Soft. Head design evolved over many iterations to increase surface area. Designs generally featured either a polygonal matrix connected to a central, tapered strut with multiple branch points or else some form of spiral (Fig. 1b). Manufacturers were able to balance sample collection (Fig. 1c), stiffness, and surface texture. Variations of a longitudinal central strut allowed for varying degrees of stability, flexibility, and impact cushioning (Fig. 1d).

### Sample and data acquisition

We collected and tested control and prototype swab pairs from 276 participants. Approximately half of the patients tested at our drive-through testing center participated. Because testing runs were batched and the COVID-19 status of participants was not known prior to testing, the number of control-positives usually exceeded the minimum requirement of 10 (range, 10-19). Total collection time was 2-3 days per prototype. The frequency of control-positive tests was 18%, generally increasing by prototype as the pandemic worsened in and around Boston.

### Comparison

All four prototypes exhibited a high degree of concordance with the control swab, with kappas of 0.88, 0.85, 0.89, and 0.88, respectively (Fig. 2a). For convenience we use the terminology of true positives, true negatives, false positives, and false negatives, with the control swab result considered the provisional gold standard. Prototypes exhibited 0-1 false positives and 1-2 false negatives. However, since control swabs are known to be an imperfect gold standard (<100% sensitivity) and because PCR positives are more likely to reflect true infection than error, false positives were interpreted as identifying missed infections; indeed, false positives were referred to clinical care teams as clinically actionable, as per IRB protocol. Of note, discordant cases were always associated with high Ct values, reflecting low viral load (Fig. 2b). For example, for Prototype 4, the control swab for one of the two false negatives had a Ct of 31.47, just shy of 31.50, our hospital’s cutoff for reportability (corresponding approximately to a single virion per mL of VTM); in addition, testing of this false negative was delayed by 16 hours because of prioritizing patient samples, which can result in decreased signal.

**Figure 2:**
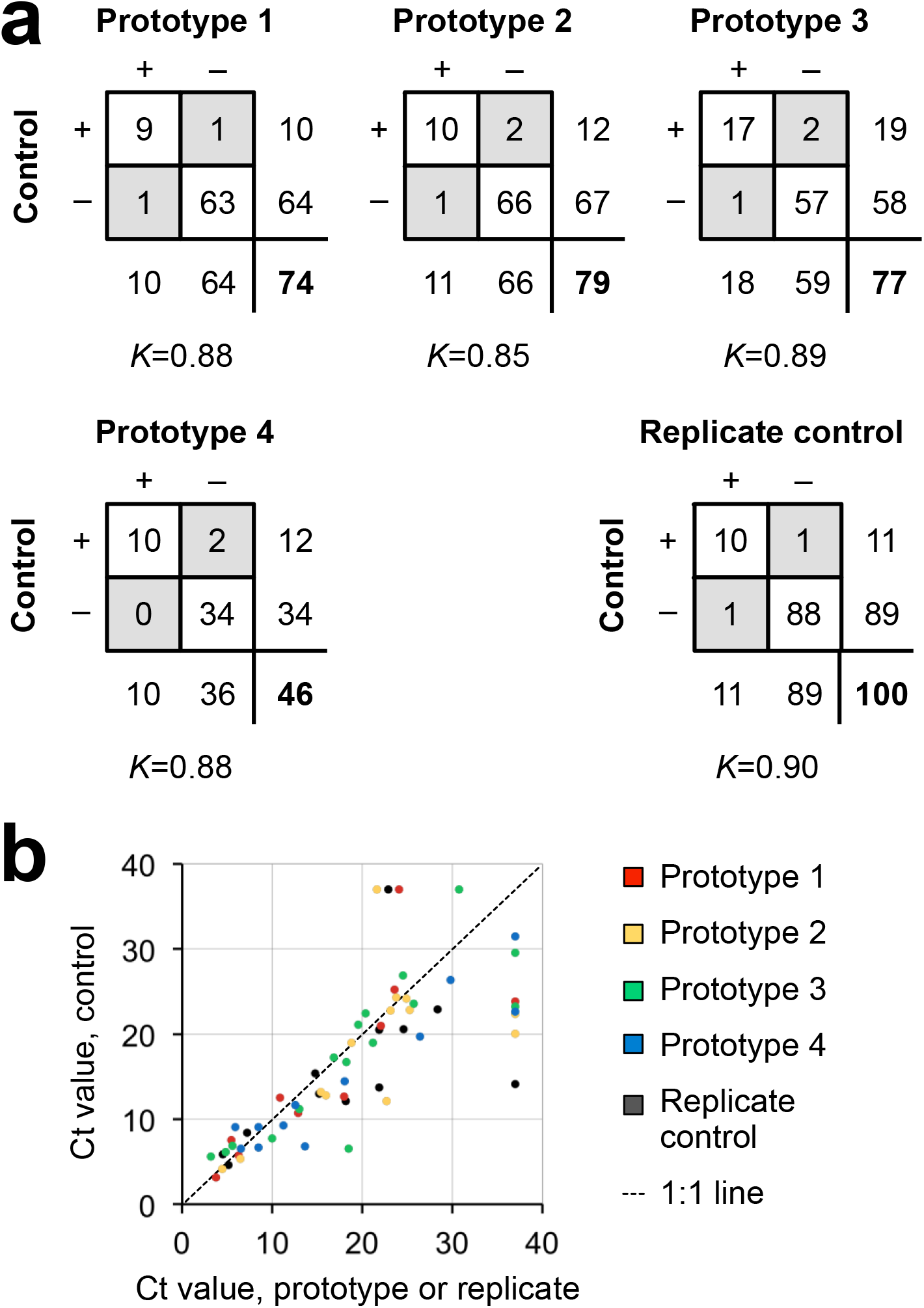
Concordance results. **(a)** 2×2 tables giving counts for each prototype vs. the control swab (first three panels) and for control vs. replicate control obtained within 24 hours on the same individual. Discordant results in gray; totals for each swab below and to the right of each box; total number of pairs in bold; *K*=Cohen’s kappa. **(b)** Scatterplot of Ct values for pairs of swabs for which at least one swab was SARS-CoV-2 positive. For discordant pairs, the negative swab was assigned a Ct value of 37 (the maximum number of cycles run).

To better assess possible performance differences between control and prototype swabs, we compared Ct values for control-prototype pairs for which at least one was positive (assigning the maximum-possible Ct to negatives; see Methods). Specifically, we asked whether the Ct values for the prototype swabs were systematically different from those of the control swabs. Systematically higher values for prototype swabs would suggest that they may underperform control swabs, notwithstanding the high kappa values. A p-value of >0.05 indicates no statistical difference. Although there were more datapoints below the 1:1 line than above it (Fig. 2b), statistical testing revealed no evidence for underperformance, with MWU p-values of 0.36, 0.26, 0.42, and 0.31 for Prototypes 1-4, respectively (Fig. 2b). This result supports the conclusion that the prototypes are non-inferior to the control.

As an additional assessment of non-inferiority, we compared the difference in Ct values observed between control and prototype swabs to the differences between replicates of control swabs. Independent of our clinical trial, there were 88 cases in which a patient, in the course of clinical care, was swabbed twice within 24 hours (mean±stdev, 15±7 hours), during the time period of our study. In 11 of these cases, at least one of the two swabs was positive for SARS-CoV-2. There were two disagreements between replicate swab tests, resulting in a kappa of 0.90, similar to what was observed in our study for each prototype (kappa=0.85–0.89). Also as in our study, the Ct values for the first swab and second swab were not significantly different (MWU p-value of 0.18). Finally, the differences between Ct values for the first and second control swabs were comparable to the differences between control and prototype swabs (MWU p-values of 0.31, 0.26, 0.47, and 0.44 for Prototypes 1-4; Fig. 2b).

### Staff and participant preferences

A written staff survey showed a preference for Prototype 4, then Prototypes 2 and 3, then Prototype 1. There was a slight preference for the control swab over Prototype 4 (Fig. 3a). In narrative feedback, Prototype 4, which underwent the largest number of revisions through our process (28), was described as comparable to the control swab (Fig. 3b).

**Figure 3:**
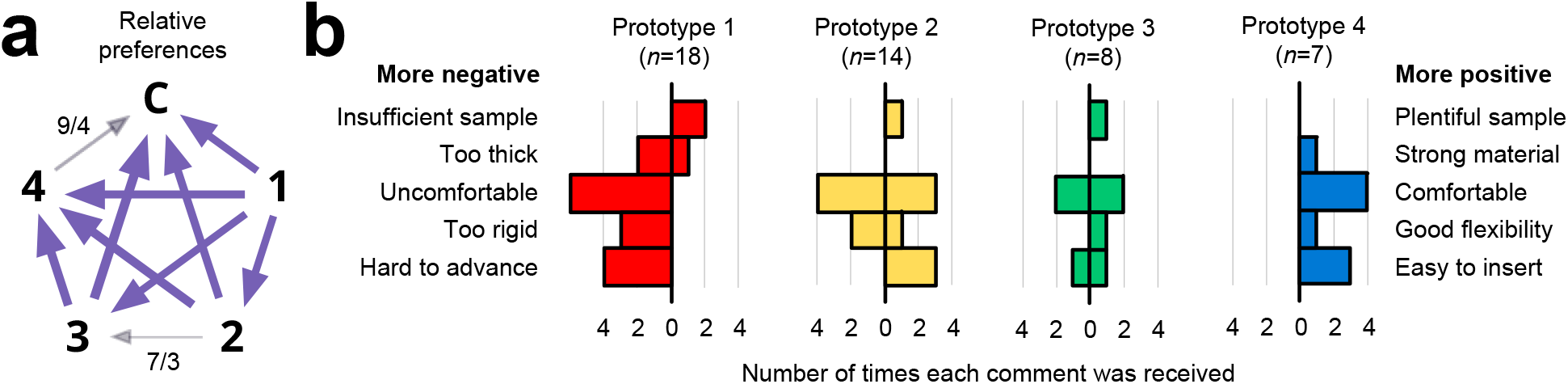
Subjective feedback. **(a)** Round-robin A-B testing of net preferences among Prototypes 1–3 (large bold numbers) and the control (“C”). Each arrow points from the less preferred to the more preferred swab. Arrow weight indicates strength of relative preference. Preferences were unanimous except where noted with numbers separated by a slash: the first number denotes the number of responses for the direction indicated by the arrowhead, while the second number denotes the number of responses that had the opposite preference. The weight of the arrow is proportional to the difference (e.g. 7-3=a net preference of 4). Unless noted, each arrow denotes 12-15 separate responses. **(b)** Number of positive and negative comments received from study staff who administered the swabs, tabulated by category. In each plot, negative feedback is to the left of the zero, while positive feedback is to the right. Bars on both the positive and negative sides of zero reflect differing opinions among study staff. *n*, total number of comments received about each prototype.

### Availability

Swabs are available to order. Details can be found on the GitHub repository in the updates at https://github.com/rarnaout/Covidswab/tree/master/BIDMC.

## Discussion

The COVID-19 pandemic has forced healthcare providers to seek alternative sources of critical materials affected by supply-chain disruptions and increases in demand. The situation has forced providers to innovate under extraordinary time-pressure. Over the course of our study we received numerous anecdotal reports of swab shortages at hospitals across the United States and in Europe, necessitating urgent stopgap solutions. Scientific literature on time-sensitive innovation suggests that open, collaborative, decentralized processes outperform closed or proprietary ones (*7–9*). Here we report the success of such a process, going from the identification of the swab crisis to multiple clinically validated prototypes capable of high-volume manufacture beginning at 22 days. Notably, none of the prototypes tested were flocked, yet their performance was statistically indistinguishable from the flocked control swab.

The urgency of the situation, the configuration of the manufacturing ecosystem, and human nature contributed to several observations and shortcomings worth mentioning. First, 3D printing has important advantages in a crisis, including the ability to iterate designs and output swabs rapidly. It remains to be seen how complementary manufacturing techniques, each with advantages and disadvantages relative to 3D printing, will contribute in a more mature market and less urgent setting. Second, in any cooperative process there is a temptation to “defect,” i.e. taking without giving back. Individuals and manufacturers may well exploit open knowledge for competitive advantage (*16*). This is a known price of openness that can disincentivize cooperation, absent social or structural mechanisms to enforce norms; managing this temptation took considerable effort by all. Third, ideally the study size would have been larger, and there would have been a better null model than replicates separated by many hours, to which to compare our results. Possible sources of variance in our study include differences in secretions or viral burden between nares and the possibility that the first (control) swab left less material for the second (prototype) when the same naris was used for both swabs. Despite these potential issues, our statistical tests supported analytical non-inferiority for all four prototypes. And fourth, we note our “round-robin” A-B testing survey was useful in summarize preferences, although the narrative comments seemed often to be more positive than the round robin suggested. A possible explanation is that the control swab was preferred in large part simply due to its being familiar, and preferred only narrowly (if often).

Like the control swab, the prototype swabs we tested can be improved upon, and manufacturers are currently doing so. The same is true for other prototypes we may test through our ongoing clinical trial. Especially in a crisis, perfect is the enemy of good enough. The pandemic continues to change quickly, and bottlenecks will likely continue to appear unpredictably. The constant requirement is the ability to respond in a timely fashion under this extraordinary pressure. We hope our experience, based on past scientific work on cooperation and innovation, will provide a useful case study for how to iterate and produce a clinically validated medical manufacture under the pressure of an ongoing pandemic, work on which others will hopefully improve as we continue to fight COVID-19 together

## Data Availability

The data supporting the findings of this study is available at https://github.com/rarnaout/Covidswab

## Acknowledgements

The authors would like to thank Abigail Bakken, Alisa Chau, Monique Cole, Zachary Fitts, Jonathan Goldie, Lan Huynh, Christina Lexinger-Holahan, Lorinda Longhi, Restituto Miranda, Pavel Gorelik, Jenny Hu, Ofer Mazor, Goutam Reddy, Karen Robinson, Stefan Riedel, Christina Yen, Summer Decker, Don Ingber, Pawan Jolly, Kit Parker, Adama Sesay, Craig Broady, John Burpo, Daniel Davis, Joe DeSimone, Annette Friskopp, Ric Fulop, Grant Michael Gonzalez, Markus Greiner, Marie Herring, Matthew Hurley, Hardik Kabaria, Shawn Patterson, David Lakatos, Ben Linville-Engler, Oren Mechanic, Richard Novak, Jifei Ou, Michael Papish, Steve Pollack, Chris Prucha, Christian Reed, Isabel Sanz, Al Siblani, Lihua Zhao, Stephanie Dominguez, Stuart Steinbock, Greg Mark, Nic Scarfo, John Bentley, Nira Pollock, Paula Watnick, Colleen Baker, Carol Daugherty. Clementina DiMonda, Virginia Dolan, Isaac Fombuh, Kylie Griffin, Amy Guadognoli, Allison Wang, Dan Eiref, Johan Sunryd, Jonathan Ford, Sherry Yu, and Kelsey Ladt for participation in the open process that made this study possible.K.E.Z. was supported by a National Institute of Allergy and Infectious Diseases training grant (T32AI007061).

K.P.S. was supported by the National Institute of Allergy and Infectious Diseases of the National Institutes of Health under award number F32AI124590.

This work was conducted with support from Harvard Catalyst | The Harvard Clinical and Translational Science Center (National Center for Advancing Translational Sciences, National Institutes of Health Award UL 1TR002541) and financial contributions from Harvard University and its affiliated academic healthcare centers. The content is solely the responsibility of the authors and does not necessarily represent the official views of Harvard Catalyst, Harvard University and its affiliated academic healthcare centers, or the National Institutes of Health.

This work was supported by Carbon3D, Envisiontec, Origin, and HP to cover the cost of the IRB and PCR testing.

The content is solely the responsibility of the authors and does not necessarily represent the official views of the National Institutes of Health.

## References

1. COVID-19 Map - Johns Hopkins Coronavirus Resource Center. https://coronavirus.jhu.edu/map.html. Accessed April 21, 2020.

2. Thomas K. Coronavirus Test Obstacles: A Shortage of Face Masks and Swabs. The New York Times. https://www.nytimes.com/2020/03/18/health/coronavirus-test-shortages-face-masks-swabs.html. Published March 18, 2020. Accessed April 13, 2020.

3. Kodzius R, Xiao K, Wu J, et al. Inhibitory effect of common microfluidic materials on PCR outcome. Sensors and Actuators B: Chemical. 2012;161(1):349–358. doi:10.1016/j.snb.2011.10.044

4. Bessetti J. An Introduction to PCR Inhibitors. 2007. https://www.promega.es/-/media/files/resources/profiles-in-dna/1001/an-introduction-to-pcr-inhibitors.pdf?la=es-es. Accessed April 13, 2020.

5. Tack P, Victor J, Gemmel P, Annemans L. 3D-printing techniques in a medical setting: a systematic literature review. BioMed Eng OnLine. 2016;15(1):115. doi:10.1186/s12938-016-0236-4

6. rarnaout. Rarnaout/Covidswab.; 2020. https://github.com/rarnaout/Covidswab. Accessed April 13, 2020.

7. Lakhani KR, Boudreau KJ, Loh P-R, et al. Prize-based contests can provide solutions to computational biology problems. Nat Biotechnol. 2013;31(2):108–111. doi:10.1038/nbt.2495

8. Lee J, Min J, Lee H. The Effect of Organizational Structure on Open Innovation: A Quadratic Equation. Procedia Computer Science. 2016;91:492–501. doi:10.1016/j.procs.2016.07.128

9. Mak RH, Endres MG, Paik JH, et al. Use of Crowd Innovation to Develop an Artificial Intelligence–Based Solution for Radiation Therapy Targeting. JAMA Oncol. 2019;5(5):654. doi:10.1001/jamaoncol.2019.0159

10. BD Gram Stain Kits and Reagents. September 2017. https://legacy.bd.com/resource.aspx?IDX=19184. Accessed April 13, 2020.

11. Preparation of Viral Transport Medium SOP# DSR-052–01. March 2020. https://www.cdc.gov/coronavirus/2019-ncov/downloads/Viral-Transport-Medium.pdf.

12. Abbott RealTime SARS-CoV-2. March 2020. https://www.molecular.abbott/sal/9N77-095_SARS-CoV-2_US_EUA_Amp_PI.pdf. Accessed April 13, 2020.

13. Miller JM, Campbell S, Loeffelholz M. Changing Swabs: To Validate or Not To Validate? J Clin Microbiol. 2013;51(11):3910–3910. doi:10.1128/JCM.02023-13

14. Kwiecien R, Kopp-Schneider A, Blettner M. Concordance Analysis. Dtsch Arztebl Int. 2011;108(30):515–521. doi:10.3238/arztebl.2011.0515

15. Mann HB, Whitney DR. On a Test of Whether one of Two Random Variables is Stochastically Larger than the Other. Ann Math Statist. 1947;18(1):50–60. doi:10.1214/aoms/1177730491

16. Fehr E, Fischbacher U. The nature of human altruism. Nature. 2003;425(6960):785–791. doi:10.1038/nature02043

